# MagSlice - Ultra Low-cost, modular and rapid 3D printed anatomical models for preoperative planning and patient education

**DOI:** 10.1101/2023.10.11.23295151

**Authors:** Harrison Tucker, Edmund Pickering, Corey Fysh, Michael Y. Chen, Nicholas J Rukin, Maria A Woodruff

## Abstract

3D printed, patient-specific anatomical models have been demonstrated as effective tools for preoperative training, especially in complex surgeries involving rare cases or unique anatomy. Current, state-of-the-art solutions are expensive, requiring 3D printers with large capital and operating costs, making them impractical for common usage and less likely to be adopted in healthcare settings. Alternatively, models fabricated using low-cost printing techniques, like fused filament fabrication (FFF), fail to achieve the utility of their more expensive counterparts. To overcome this challenge, we have developed an innovative approach using FFF that is able to partition the model into separate segments, held together with magnetic fasteners, to unveil the internal anatomy of the model thereby overcoming the challenges of low-cost printing methods and providing equivalent efficacy to the state-of-the-art. Notably, our models can be printed significantly faster and cheaper than other published models and can be viewed holistically as one model, or can be physically expanded into slices (like a deck of cards) whilst maintaining the entire model integrity. By dramatically reducing cost and time, our solution unlocks the potential of 3D printed patient specific models, making these practical for hospital usage.

## 1. Introduction

Patient-specific anatomical models improve outcomes in complex surgical procedures by offering a physical representation of the anatomy, allowing clinicians to better communicate with patients [1-6]. Surgical models of the kidney are ideal for 3D printing to aid surgical planning. With smaller lesions been treated with minimally invasive surgery such as partial nephrectomy, the ability to clearly demonstrate the anatomical relationship of the kidney tumor to the renal vasculature and collecting system aids the surgeon with the planned procedure [3]. However, most models are fabricated on printers with large capital and material costs which limit adoption [7].

Fused filament fabrication (FFF) is a low-cost printing method but is limited for surgical planning models as it cannot print complex 3D geometries without support materials and it lacks transparent materials options [2, 8]. These are desired in surgical planning models as they allow the surgeon to view the internal anatomy of the organ. FFF is also time intensive for printing surgical models at scale [7].

To overcome these limitations, we developed an innovative approach to 3D printing patient-specific anatomical models based on low-cost FFF. Our approach enables both the printing of complex 3D organs and the visualization of the internal structure, by breaking the model into multiple segments (Fig. 1e) which are magnetically attached (see Supplementary Video 1). A further advantage of this approach is that users can interact with a physical representation of the model. By pulling the model apart, internal structures can be physically inspected in 3D without needing to refer to 2D medical imaging software. Our method uses 80% less in material costs compared to other published works and is significantly faster than other manufacturing methods [3, 9].

**Fig. 1.**
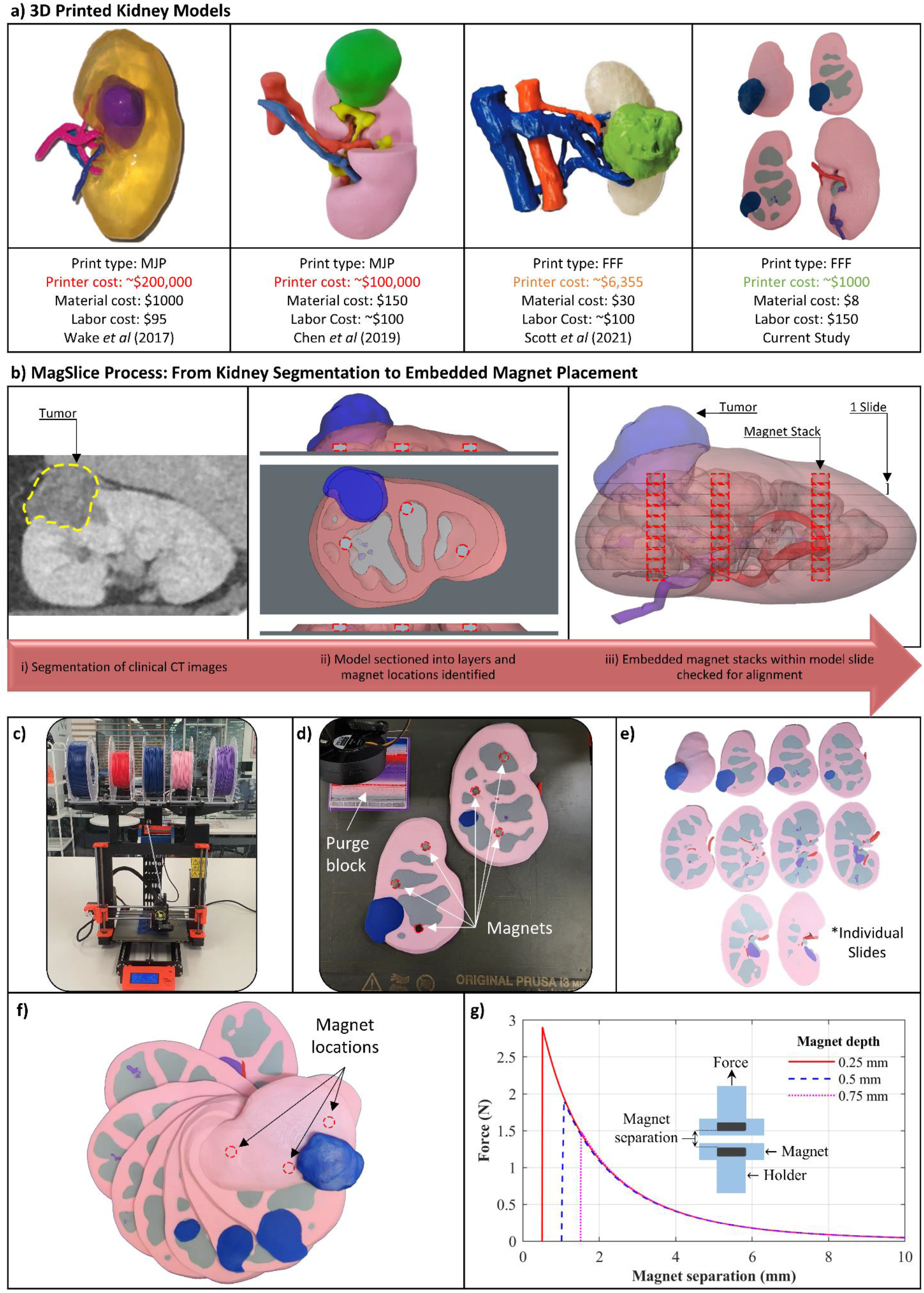
(**a**) comparison of different 3DP anatomical kidney models [4, 8, 9]. (**b**) Low-cost anatomical model pipeline: Magslice Process. (**b.i**) Clinical CT scan of kidney viewed in Materialise Mimics. (**b.ii**) Segmented model identifying proposed magnet locations within 3 slides of the kidney model. (**b.iii**) Model of kidney showing cutting planes and magnet positions. (**c**) Prusa MK3S with MMU2S used to print this model. (**d**) 3D print paused on specified layer before magnets are embedded. (**e**) Final product showing individually printed sections. (**f**) Final printed model with each layer splayed apart. (**g**) Mechanical testing of magnet pull force.

## 2 Methodology

The model fabrication workflow is detailed in Figure 1b, using a de-identified patient scan of a renal cell carcinoma (RCC) as a test case, provided by Royal Brisbane and Women’s Hospital (human research ethics committee study reference: LNR/QRBW/52822). Clinical computed-tomography (CT) scans (Helical CT, 1 mm slice thickness, contrast enhanced) of kidneys with renal cell carcinoma were segmented in Mimics v24 (Materialise, Leuven, Belgium) using thresholding tools to identify the renal artery (red), ureter (purple), renal medulla (grey) tumor (blue) and remaining parenchymal tissue (pink) (see Fig. 1b.ii) [10]. The model was imported in 3-matic v16 (Materialise, Leuven, Belgium) and cutting planes (i.e., those cross-sections which are desired to be viewed) were identified (Fig. 1b.iii). The model was split along the planes into 10 sections, with each section being 3.8 mm thick to encapsulate magnets. Cut outs were created to embed the magnets (Huifung, AliExpress (Shenzhen, Guangdong, China), 6 mm x 3 mm, material: neodymium, grade: n35, unit cost: $0.07) (Fig. 1b.iii). These were embedded 0.2 mm in both faces of the model. Boolean subtraction was used to create a cavity for the magnets to be placed. This process was repeated for each section of the model that required magnets, taking care to ensure the magnets aligned for each section, ensuring the final geometry can correctly orientate. A fully assembled model, pre-print, is shown in Figure 1b.iii.

Each section was exported as a 3MF file to slicing software (PrusaSlicer, version 2.3.3) and prepared for printing on a Prusa MK3S+ printer with a multi material upgrade 2S (MMU2S) (Prusa Research, Prague, Czech Republic) (Fig. 1c). Models were aligned on the print bed, and fabricated with polylactic acid (PLA) filament (Filaform, Adelaide, Australia) with colors assigned for different regions and a purge between print color transitions to prevent color bleed. The print was automatically paused on selected layers, allowing magnet placement in each cavity. Figure 1d shows a paused print with magnets inserted.

To assess magnet pull strength, specimens were fabricated with magnets embedded at 0.25 mm, 0.5 mm and 0.75 mm depths and mechanically loaded (Instron 5544A, 100 N loadcell, 10 mm/min load rate). Tests were in triplicate with results averaged (Fig. 1g).

## 3. Results

The fabricated model is shown in Figure 1f, with a video of the model use in Supplementary Video 1. The total fabrication time, from CT to printed model, was 29 hrs (5 hrs segmentation and model preparation, 24 hrs print time), and required 342 g of filament. The total estimated cost was $158 (considering $150 for labor and $0.023/g for filament, costs reported in USD).

## 4. Discussion

Figure 1a shows examples of published renal models. These often use the MultiJet printing (MJP) process, with a capital cost in excess of $40,000 per printer [7] and material costs between $150 [9] to $1,000 per model [4]. Alternatively, our method uses low-cost FFF, with the printer costing $799 (Prusa i3 MK3S+, Prusa Research, Prague, Czech Republic) and the Prusa i3 MMU2S upgrade kit costing $299.

Other major costs include material and labor. Material costs in MJP can be large, with an average kidney model costing upwards of $100 per model. (Average male kidney volume 203 ml, MJP material ∼$0.67/g). In the work presented here, through optimizing print and purge parameters, material costs are negligible in our model, totaling $8. Labor associated with segmentation and preparing the print are similar across printing technologies, and primarily depend on operator experience and speed. In our work, this was reduced to $150, making our total model cost of $158 per unit.

Common advantages of MJP include the use of transparent material (revealing the internal geometry) and dissolvable support material allowing detailed features to be easily cleaned [3, 7]. Other work has investigated low cost FFF methods [7, 8], but have not been able to create models of equivalent utility to the state-of-the-art. For example, Scott et al. [8], demonstrated a low-cost FFF kidney model which separated into two parts to make-visible a tumor. However, this approach does not fully reveal the internal anatomy of the kidney. Our new *Magslice* method overcomes these challenges by creating multiple thin sections, held by magnetic clasps. This allows the user to ‘open’ the model and reveal the internal geometry, allowing visualization of the model at 3.8 mm increments.

## 5. Conclusion

3D printed patient specific preoperative planning models are a valuable emerging tool. These are limited by large capital and operation costs. Our work overcomes these challenges by developing methods for producing models of equivalent utility to the state-of-the-art, utilizing the ultra-low cost FFF method. By dramatically reducing the cost, our method unlocks the potential of 3D printed patient specific preoperative planning models.

## Data Availability

All data produced in the present work are contained in the manuscript

## Abbreviations

RCC: Renal cell carcinoma
3MF: 3D Manufacturing Format
MMU: Multi material upgrade
FFF: Fused filament fabrication
MJP: MultiJet printing
CT: Computed tomography
PLA: Polylactic acid

## Acknowledgements

Dr Pickering is supported by a Centre for Biomedical Technologies research fellowship. The authors would like to acknowledge the Royal Brisbane and Women’s Hospital for providing RCC scans.

